# Critical care demand and intensive care supply for patients in Japan with COVID-19 at the time of the state of emergency declaration in April 2020: a descriptive analysis

**DOI:** 10.1101/2020.06.20.20136150

**Authors:** Yosuke Fujii, Kiichi Hirota

## Abstract

**Background:** The coronavirus disease 2019 (COVID-19) pandemic overwhelms Japan’s intensive care capacity. This study aimed to determine the number of patients with COVID-19 who required intensive care and to compare the numbers with Japan’s intensive care capacity.

**Methods:** Publically available datasets were used to obtain the number of confirmed patients with COVID-19 undergoing mechanical ventilation and extracorporeal membrane oxygenation (ECMO) between February 15 and July 19, 2020, to determine and compare intensive care unit (ICU) and attending bed needs for patients with COVID-19, and to estimate peak ICU demands in Japan.

**Results:** During the epidemic peak in late April, 11443 patients (1.03/10000 adults) had been infected, 373 patients (0.034/10000 adults) were in ICU, 312 patients (0.028/10000 adults) were receiving mechanical ventilation, and 62 patients (0.0056/10000 adults) were under ECMO per day. At the peak of the epidemic, the number of infected patients was 651% of designated beds, and the number of patients requiring intensive care was 6.0% of ICU beds, 19.1% of board-certified intensivists, and 106% of designated medical institutions in Japan.

**Conclusions:** The number of critically ill patients with COVID-19 continued to rise during the pandemic, exceeding the number of designated beds but not exceeding ICU capacity.

## Introduction

A novel coronavirus, namely, severe acute respiratory syndrome coronavirus 2 (SARS-CoV-2), was first reported in Wuhan, China, in December 2019, and the number of newly reported cases has continued to increase globally. As of September 2020, >34 million cases had been reported worldwide and >1000000 people had died [1], with >84000 reported cases and >1500 deaths recorded in Japan. On January 30, 2020, the World Health Organization (WHO) declared the epidemic to be a “public health emergency of international concern”. On April 7, 2020, Japan declared a state of emergency in seven major central cities, which then was extended to all 47 prefectures on April 16, 2020 [2].

COVID-19 affects the respiratory system and patients infected with SARS-CoV-2 suffer from respiratory syndromes [3]. Although approximately 70-80% of patients with COVID-19 have mild to moderate symptoms and recover, 5-20% of patients develop severe respiratory failure and may require admission to an intensive care unit (ICU) [3-5].

Symptomatic treatment for severe respiratory failure has involved the use of mechanical ventilation and extracorporeal membrane oxygenation (ECMO) [5-9] although there are candidate drugs available to treat severe patients with COVID-19 as of September 2020 [10,11]; remdesivir [12,13] and dexamethasone [14]. One study that analyzed clinical cases of COVID-19 reported a better prognosis in patients with a mild infection, whereas the prognosis for patients with severe and critical infections is poor. However, patients presenting with mild infection are less obvious and are less easily located or identified [15]. A previous observational study reported that approximately 50% of the patients admitted to ICU required ventilation support and, of these, one in four patients required considerably greater levels of intensive life support [15]. A cohort study found that approximately 8% of patients with COVID-19 were critically ill [16]. One case series reported that, of 18 patients who required intubation, 1 patient required ECMO and 5 patients died [17]. It has been estimated that, despite intensive life support, approximately 3-4% of all infected patients are likely to die due to SARS-CoV-2 infection [3,18].

The importance of ICUs during the COVID-19 pandemic has been well established; however, ICU capacity concerning expected numbers of patients with COVID-19 requiring intensive care is unknown. During the COVID-19 epidemic, there has been a dramatic surge in the number of critically ill patients with an urgent need for ICU admission [19], as occurred in other natural disasters [20,21]. An analysis of the epidemic peak in Wuhan indicated that the number of severely and critically ill adult patients requiring intensive care was estimated to be 12.2/10000 and 2.6/10000, respectively [22]. However, this situation differs between countries, and specific data sourced in terms of Japanese circumstances are needed [23]. This study aimed to determine the number of critically ill patients who required ICU admission, including the number of patients requiring mechanical ventilation and ECMO, and to compare these patient numbers with ICU capacity in Japan at nationwide and regional levels.

## Materials and Methods

### Ethical approval

Institutional review board approval and patient informed consent were not required for this study, as this study did not involve human participant research or any interventions.

### Infectious disease data source

We obtained data concerning numbers of infected, recovered, and dead patients due to COVID-19 in Japan from a publicly available website [24]. The number of patients who required mechanical ventilation and ECMO support was extracted from the Japanese Society of Intensive Care Medicine (JSICM) website [23,25]. This database accumulates data using a CRoss Icu Searchable Information System (CRISIS). More than 570 facilities across Japan currently participate in this CRISIS, centering on facilities certified by the JSICM and facilities designated by the Japanese Society of Emergency Medicine, with a total of 5500 ICU beds, covering 80% of the total ICU beds in Japan [26]. This database provides information concerning the number of patients under mechanical ventilation, under ECMO, the number of patients in recovery from ECMO, and the number of patients who died during ECMO.

### Intensive care unit capacity data source

We extracted the number of ICU beds (n = 7109) and board-certified intensivists (n = 1957) from the JSICM website. This website has also reported the estimated number of mechanical ventilators for adults (n = 30091) and ECMO machines (n = 2208) in Japan (table 1) [27].

**Table 1.** Critical care capacity in Japan and seven regions. The raw count and the proportional count per 10000 adults in parenthesis were listed. *: 100000 adults. Category II; designated medical institutions for Category II infectious diseases, TB; tuberculosis, ECMO; extracorporeal membrane oxygenation.

|  | Hokkaido | Tohoku | Kanto-Koshinetsu | Tokai-Hokuriku | Kansai | Chugoku-Shikoku | Kyusyu-Okinawa | Japan |
| --- | --- | --- | --- | --- | --- | --- | --- | --- |
| Prefecture | 1 | 6 | 10 | 7 | 6 | 9 | 8 | 47 |
| Population<br>( 10<sup>5</sup> adults) | 468.5 | 769.8 | 4287 | 1565.3 | 1805.4 | 966.6 | 1233.4 | 11075.4 |
| Category II beds | 92<br>(0.02) | 171<br>(0.022) | 531<br>(0.012) | 220<br>(0.014) | 238<br>(0.013) | 206<br>(0.021) | 300<br>(0.024) | 1758<br>(0.016) |
| Category II<br>+ TB beds | 153<br>(0.033) | 261<br>(0.034) | 937<br>(0.022) | 513<br>(0.033) | 616<br>(0.034) | 397<br>(0.041) | 625<br>(0.051) | 3502<br>(0.032) |
| Intensivist | 75<br>(0.016) | 95<br>(0.012) | 727<br>(0.017) | 233<br>(0.015) | 368<br>(0.02) | 228<br>(0.024) | 231<br>(0.019) | 1957<br>(0.018) |
| Category II<br>institution | 24<br>(0.005) | 40<br>(0.005) | 91<br>(0.002) | 46<br>(0.003) | 39<br>(0.002) | 42<br>(0.004) | 69<br>(0.006) | 351<br>(0.003) |
| ECMO project<br>hospital* | 4<br>(0.0085) | 8<br>(0.0104) | 35<br>(0.0082) | 11<br>(0.007) | 24<br>(0.0133) | 6<br>(0.0062) | 12<br>(0.0097) | 100<br>(0.009) |

In Japan, COVID-19 was designated as an infectious disease under the Infectious Diseases Control Law on January 28, 2020. Patients with COVID-19 were admitted and transferred to designated medical institutions responsible for the hospitalization of patients with a Category II infectious disease (especially tuberculosis) or a novel influenza infection. Although clear evidence of person-to-person airborne transmission of SARS-CoV-2 has not been established, an airborne component of transmission is likely, based on other respiratory viruses such as severe acute respiratory syndrome (SARS), Middle East respiratory syndrome (MERS), and influenza [28-31] and its presence in aerosol has been confirmed at RNA level [32,33]. Many infectious control guidelines recommend that healthcare workers be protected through the use of personal protective equipment [34-36]. Suspicious airborne infectious routes make patient care difficult and complex, requiring the management of the patient using airborne precautions [37] or management of the patient in a depressurized area [36]. A patient management ratio of one critically ill patient to one depressurized area per hospital has been used, assuming that not all infected patients could be treated in ICU beds. In Japan, there are 1758 beds designated for patients with a Category II infectious disease, 3502 beds allocated for patients with tuberculosis, and 351 designated medical institutions for patients with Category II infectious diseases (table 1) [38].

ECMO is a highly integrated critical care treatment that requires multidisciplinary team collaboration [39,40]. It is also assumed that not all of the ECMO machines in Japan could be allocated to manage critically ill patients with COVID-19. In Japan, an organized ECMO-based respiratory program, namely, the ECMO project [41], comprises a consortium of specialized institutions for ECMO involving the participation of 100 hospitals in this project. We assumed these hospitals were the institutions specially dedicated to treating critically ill patients with COVID-19 who required treatment using ECMO machines (table 1) [41].

### Board-certified intensivists and patient numbers in ICUs in seven regions in Japan according to the JSICM and CRISIS

Data concerning population numbers in Japan in 2019 at nationwide and regional levels and the proportion of adults aged ≥15 years were retrieved from the Statistics Bureau of Japan [42]. The national population was 1.26 ×10^8^ and the proportion of adults was 87.9%. To determine regional differences, we analyzed the demand and supply of ICUs in the 47 prefectures. The JSICM and CRISIS have reported the number of board-certified intensivists and the number of patients in ICU in seven regions in Japan, namely, Hokkaido, Tohoku, Kanto-Koshinetsu, Tokai-Hokuriku, Kansai, Chugoku-Shikoku, and Kyusyu-Okinawa (figure 1). Data concerning the 47 prefectures were then integrated according to these seven regions (table 1).

**Figure 1.**
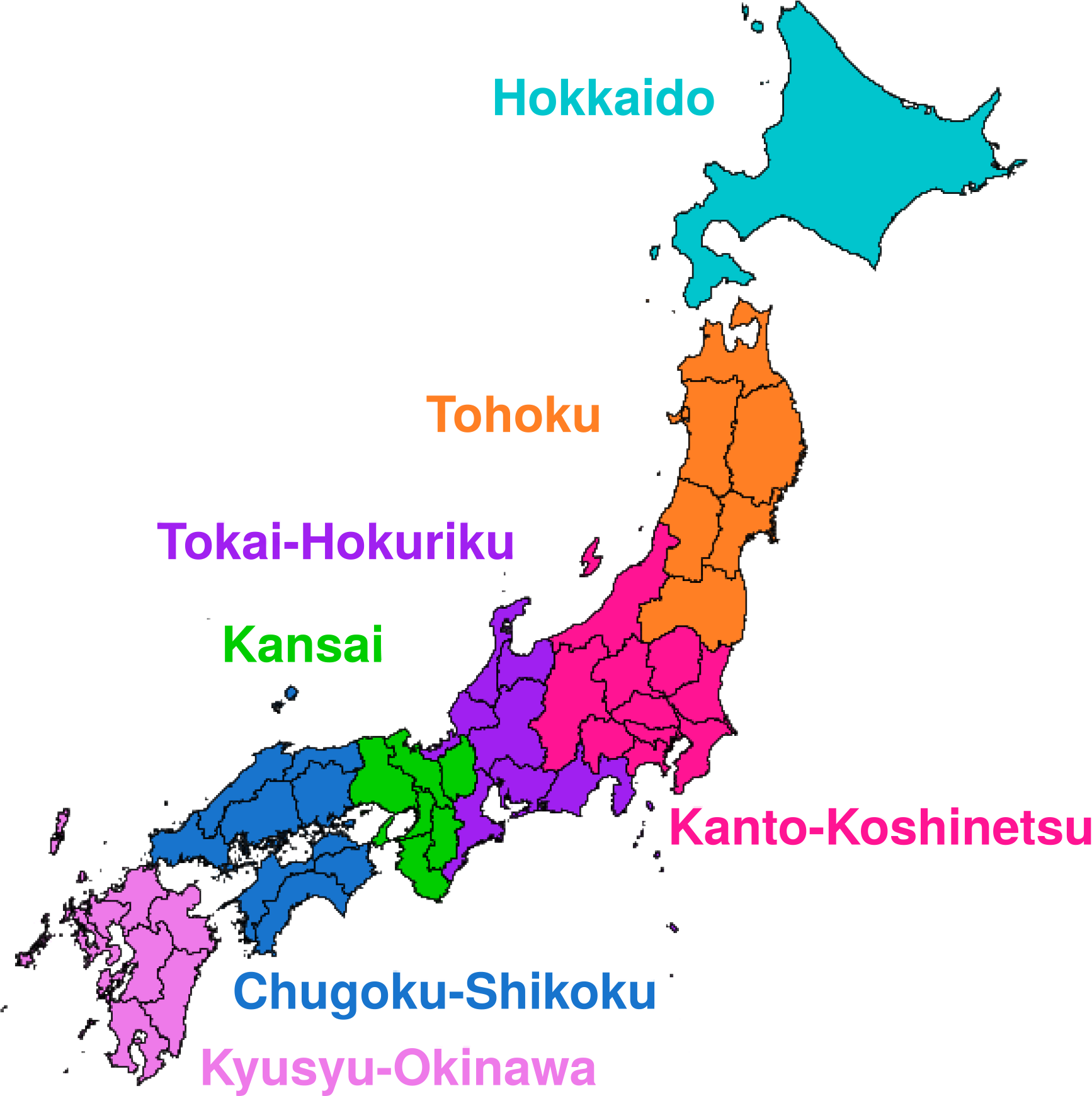
Seven regions in Japan defined by the Japanese Society of Intensive Care Medicine. Seven regions are Hokkaido, Tohoku (Aomori, Iwate, Akita, Miyagi, Yamagata, and Fukushima), Kanto-Koshinetsu (Ibaraki, Tochigi, Gumma, Saitama, Chiba, Tokyo, Kanagawa, Niigata, Yamanashi, and Nagano), Tokai-Hokuriku (Toyama, Ishikawa, Fukui, Gifu, Shizuoka, Aichi, and Mie), Kansai (Shiga, Kyoto, Osaka, Hyogo, Nara, and Wakayama), Chugoku-Shikoku (Tottori, Shimane, Okayama, Hiroshima, Yamaguchi, Tokushima, Kagawa, Ehime, and Kochi), and Kyushu-Okinawa (Fukuoka, Saga, Nagasaki, Kumamoto, Oita, Miyazaki, Kagoshima, and Okinawa).

### Statistical analysis

The total number of patient days in ICU was summed to estimate the total number of ICU days and the raw number of patients in ICU on each day was plotted [22]. The notation “critical case” denoted the total number of patients on mechanical ventilator and ECMO. The proportion of critically ill patients per 10000 adults was estimated based on the number presented in the infectious diseases and ICU capacity data source sections.

## Results

COVID-19 accounted for a total of 21705 ICU days, 18495 mechanical ventilation days, and 3210 ECMO days between January 15, 2020, and July 19, 2020, with a median (interquartile range [IQR]) of 96 (68-197) patients in ICU, 83 (58-174) patients on mechanical ventilation, and 15 (11-27) patients on ECMO machines each day during these 156 days (figure 2 and table 2). During the peak of the epidemic in late April 2020, 11443 patients (1.03 per 10000 adults) had been infected with SARS-CoV-2 and, of these, 373 patients (0.034 per 10000 adults) were admitted to ICU, 312 patients (0.028 per 10000 adults) were treated using mechanical ventilation, and 62 patients (0.0056 per 10000 adults) were treated using ECMO machines per day.

**Table 2.** ICU-, ventilation-, and ECMO-day for seven regions and Japan. The total days and 0.25, 0.5, and 0.75 quartile days in brackets were shown. ICU; intensive care unit, ECMO; extracorporeal membrane oxygenation.

|  | Hokkaido | Tohoku | Kanto-Koshinetsu | Tokai-Hokuriku | Kansai | Chugoku-Shikoku | Kyusyu-Okinawa | Japan |
| --- | --- | --- | --- | --- | --- | --- | --- | --- |
| critical case | 1766<br>[8, 10, 13] | 200<br>[0, 1, 2] | 11805<br>[40, 51, 106] | 1318<br>[5, 8, 12] | 4587<br>[6, 17, 48] | 367<br>[0, 2, 3] | 1662<br>[1, 10, 17] | 21705<br>[68, 96, 197] |
| ventilation | 1547<br>[7, 9, 12] | 170<br>[0, 1, 1] | 9820<br>[30, 44, 93] | 1102<br>[4, 7, 10] | 4188<br>[6, 14, 44] | 358<br>[0, 2, 3] | 1310<br>[1, 9, 14] | 18495<br>[58, 83, 174] |
| ECMO | 219<br>[1, 1, 2] | 30<br>[0, 0, 0] | 1985<br>[7, 10, 16] | 216<br>[1, 1, 2] | 399<br>[0, 0, 4] | 9<br>[0, 0, 0] | 352<br>[0, 2, 3] | 3210<br>[11, 15, 27] |

**Figure 2.**
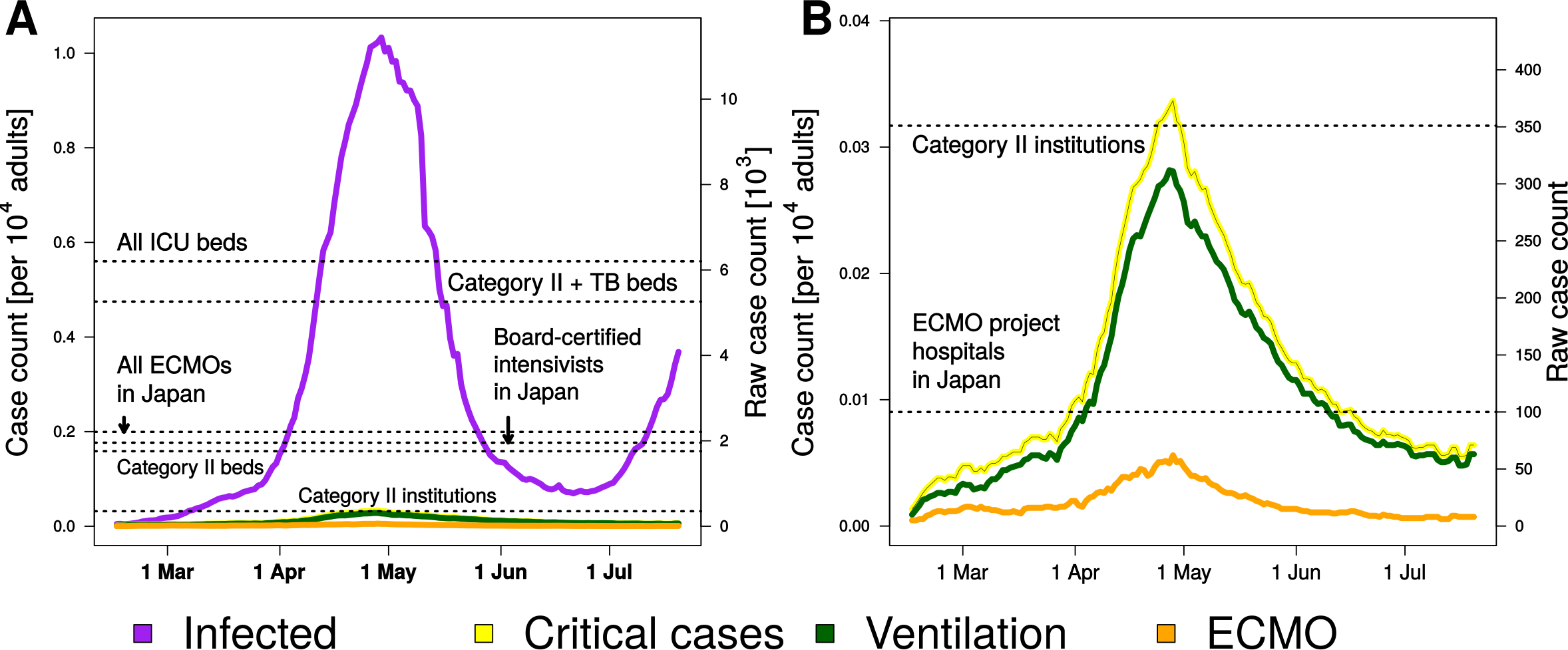
The burden of critical coronavirus disease during the epidemic in Japan. (A) The number of infected and critically ill patients per 1000 adults. Critically ill patients were the sum of the number of patients under mechanical ventilation and ECMO. The Japanese government also declared “The State of Emergency” on 7 April 2020 for major central cities with a cumulative 4257 confirmed cases and 93 deaths among patients with COVID-19 and it spread to all of 47 prefectures on 16 April 2020. (B) The magnified version of figure A. Category II; designated medical institutions for Category II infectious diseases, TB; tuberculosis, ECMO; extracorporeal membrane oxygenation.

The number of infected patients at the peak of the epidemic was 651% of total designated beds for Category II infectious disease and 218% of total Category II with tuberculosis beds, and the number of patients requiring intensive care at the peak of the epidemic was 6.0% of total ICU beds in Japan, 19.1% of total board-certified intensivists in Japan, and 106% of total Category II institutions. The number of patients requiring ECMO at the peak of the epidemic was 3.2% of total board-certified intensivists in Japan, 17.7% of total Category II institutions, and 62% of total hospitals participating in ECMO project in Japan, respectively (figure 2 and table 3).

**Table 3.**
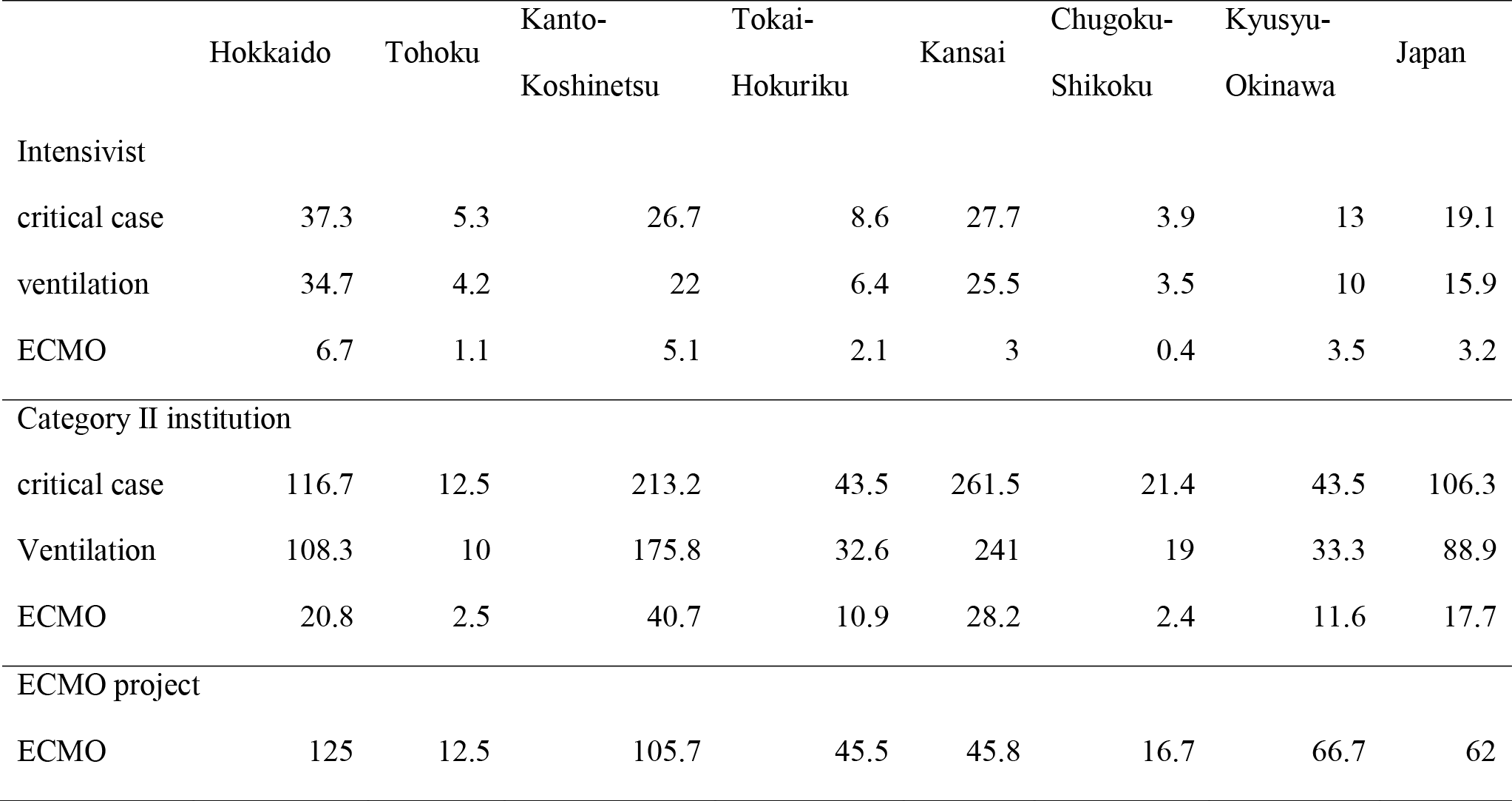
The percentage of the number of patients requiring critical care, mechanical ventilation, and ECMO to the number of board-certified intensivists, Category II institutions, and ECMO project hospitals for each region in Japan at the peak of the epidemic. ICU; intensive care unit, Category II; a designated medical institution for Category II infectious diseases, TB; tuberculosis, ECMO; extracorporeal membrane oxygenation.

|  | Hokkaido | Tohoku | Kanto-Koshinetsu | Tokai-Hokuriku | Kansai | Chugoku-Shikoku | Kyusyu-Okinawa | Japan |
| --- | --- | --- | --- | --- | --- | --- | --- | --- |
| Intensivist |  |  |  |  |  |  |  |  |
| critical case | 37.3 | 5.3 | 26.7 | 8.6 | 27.7 | 3.9 | 13 | 19.1 |
| ventilation | 34.7 | 4.2 | 22 | 6.4 | 25.5 | 3.5 | 10 | 15.9 |
| ECMO | 6.7 | 1.1 | 5.1 | 2.1 | 3 | 0.4 | 3.5 | 3.2 |
| Category II institution |  |  |  |  |  |  |  |  |
| critical case | 116.7 | 12.5 | 213.2 | 43.5 | 261.5 | 21.4 | 43.5 | 106.3 |
| Ventilation | 108.3 | 10 | 175.8 | 32.6 | 241 | 19 | 33.3 | 88.9 |
| ECMO | 20.8 | 2.5 | 40.7 | 10.9 | 28.2 | 2.4 | 11.6 | 17.7 |
| ECMO project |  |  |  |  |  |  |  |  |
| ECMO | 125 | 12.5 | 105.7 | 45.5 | 45.8 | 16.7 | 66.7 | 62 |

The number of total ICU days was 1766 (IQR, 8-13) in Hokkaido, 200 (0-2) days in Tohoku, 11805 (40-106) days in Kanto-Koshinetsu, 1318 (5-12) days in Tokai-Hokuriku, 4587 (6-48) days in Kansai, 367 (0-3) days in Chugoku-Shikoku, and 1662 (1-17) days in Kyusyu-Okinawa (figure 3 and table 2).

**Figure 3.**
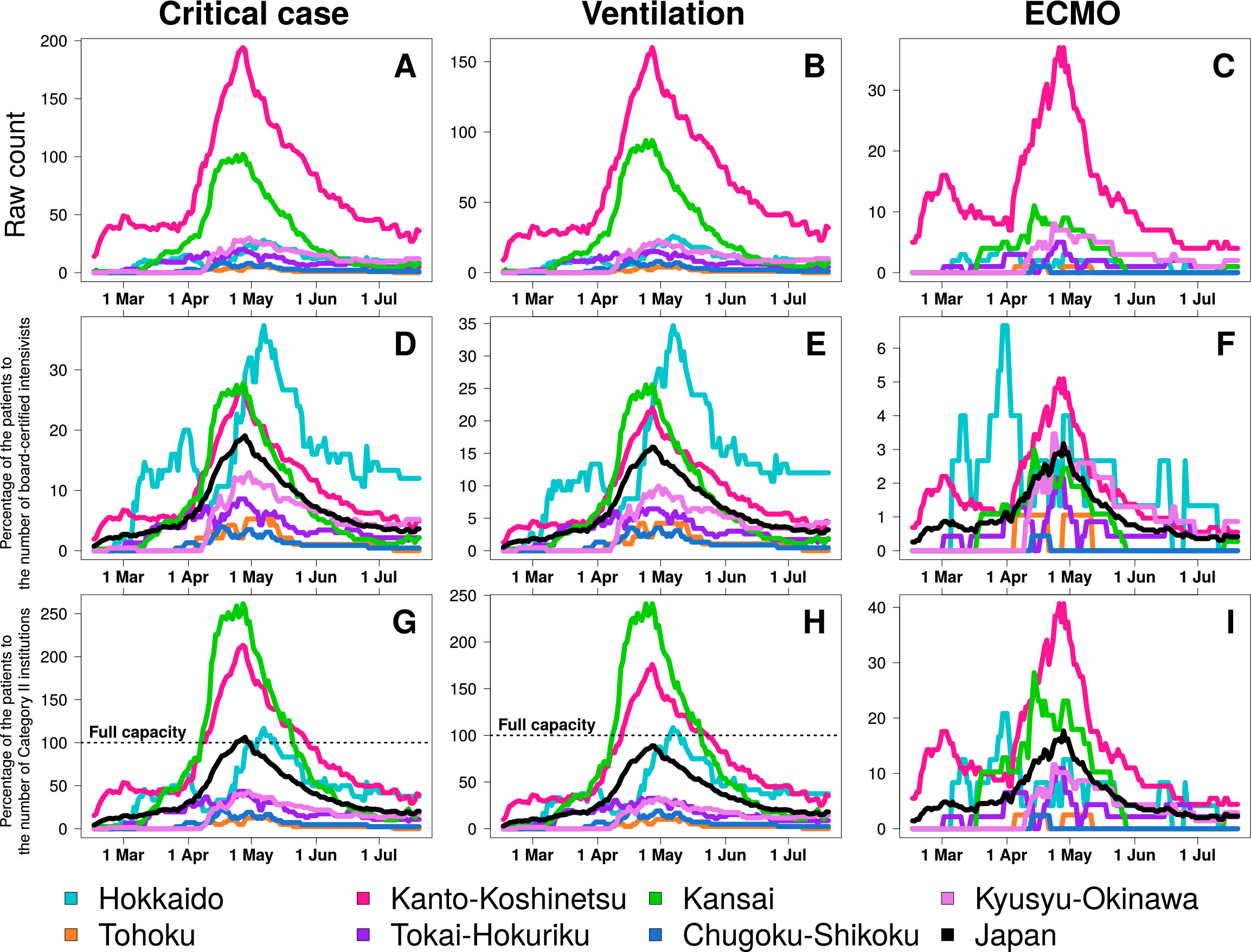
The burden of critical coronavirus disease during the epidemic in seven regions defined by the Japanese Society of Intensive Care Medicine in Japan. Raw count of COVID-19 patients under critical care (A), mechanical ventilation (B), and ECMO (C), percentage of patients to the number of board-certified intensivists for critical care (D), mechanical ventilation (E), and ECMO (F), and percentage of patients to the number of Category II institutions for critical care (G), mechanical ventilation (H), and ECMO (I), respectively. Category II; designated medical institutions for Category II infectious diseases, ECMO; extracorporeal membrane oxygenation.

ICU demand compared with the proportion of board-certified intensivists greatly increased in Hokkaido (37.3% of total board-certified intensivists in Hokkaido), in Kanto-Koshinetsu (26.7%), and Kansai (27.7%) compared with the overall demand in Japan (19.1%). ICU and mechanical ventilation demand overwhelmed the number of Category II institutions in Hokkaido (117% of total Category II institutions of ICU and 108% of total Category II institutions for mechanical ventilation), in Kanto-Koshinetsu (213% for ICU and 176% for mechanical ventilation), and Kansai (262% for ICU and 241% for mechanical ventilation) (table 3). The number of patients requiring ECMO compared with the number of ECMO project hospitals was 125% in Hokkaido and 106% in Kanto-Koshinetsu (figure 4 and table 3).

**Figure 4.**
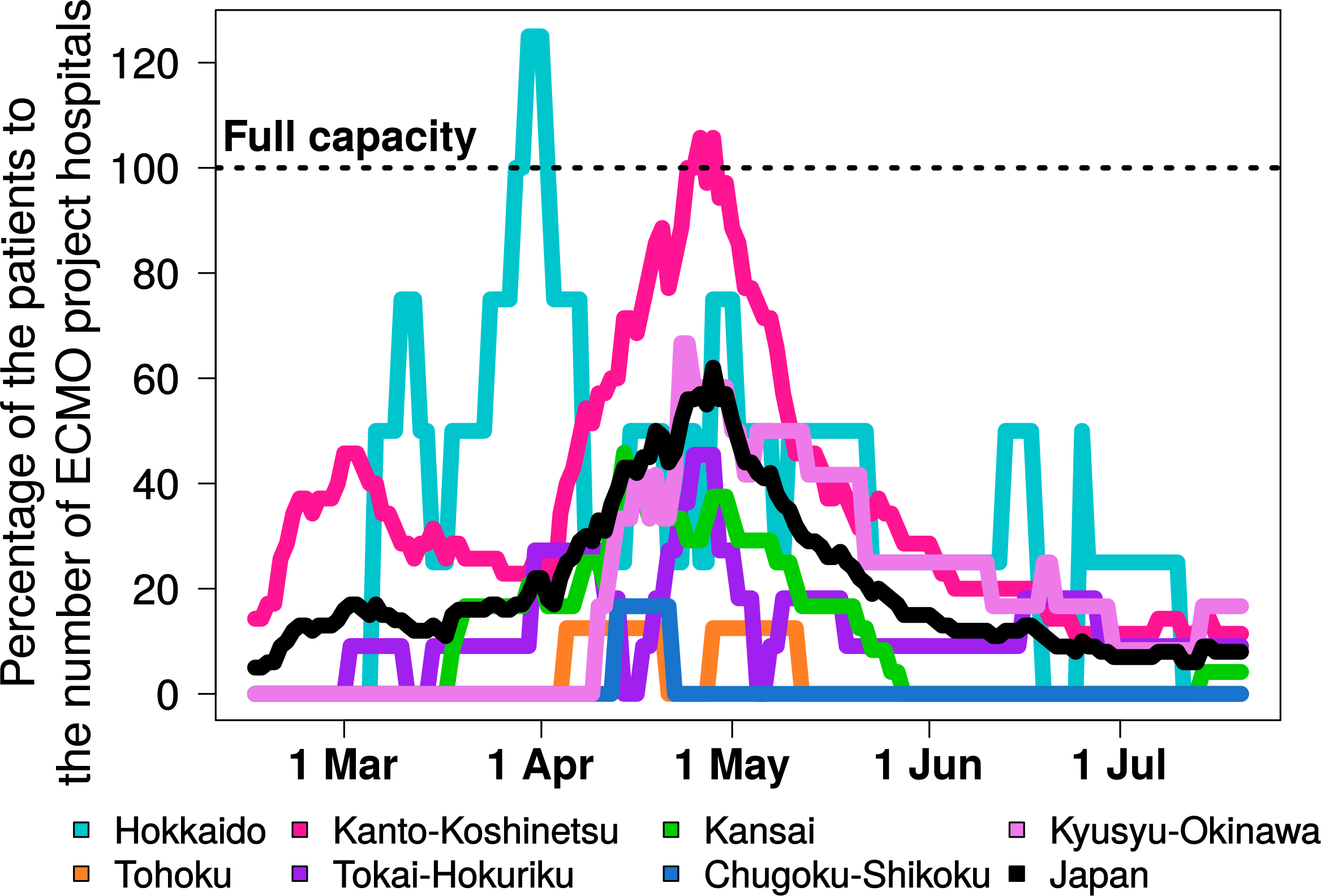
The burden of critical coronavirus disease requiring ECMO to the number of ECMO project hospitals during the epidemic in seven regions defined by the Japanese Society of Intensive Care Medicine in Japan. ECMO; extracorporeal membrane oxygenation.

## Discussion

The number of new patients diagnosed with COVID-19 continues to increase globally. Approximately 70-80% of patients infected with SARS-CoV-2 show mild to moderate respiratory symptoms; however, between 5% and 32% of these patients may develop severe symptoms and require ICU admission, of whom between 3% and 22% may require mechanical ventilation and between 0.5% and 5% may require oxygenation with ECMO [3,4,15,37,43-45]. Depending on the efficacy of infection control strategies, a shortage of ICU ventilators may or may not eventuate [41].

Exceeding hospital bed capacity would likely increase the community spread of SARS-CoV-2 and lead to decreased quality of healthcare and intensive care [22]. In China, national lockdown, quarantine, and social distancing measures were adopted to control the spread of infection, with >10000 beds designated for the isolation and treatment of patients with confirmed COVID-19 by early February 2020 [22]. Despite these efforts to control the spread of infection, this did not lead to an immediate downturn in demand for hospitalization in critical care units and ICUs in China [22], and this situation has been similar in Japan.

Our results indicated a critical need to isolate and treat patients with COVID-19; however, while there remained a small margin for critically ill patients requiring mechanical ventilation and ECMO in terms of ICU capacity, this situation was sub-optimal. Intensivists do not necessarily attend to all critically ill patients and have been reported to provide care to only 37% of all ICU patients [46]. In the United States, one study reported that only approximately 30% of intensive care units were staffed by dedicated intensivists [47]. Due to different health care systems, statistics from the United States model could not be simply extrapolated to Japan, but we have assumed that these percentages are likely to be the same or no greater in Japan. The number of critically ill patients in ICU at the peak of the epidemic accounted for 19% of total board-certified intensivists in Japan, which challenged Japan’s ICU capacity.

Our findings also showed a regional bias in terms of ICUs being overwhelmed in Japan. Kanto-Koshinetsu and Kansai are the biggest regions in Japan. The Kanto-Koshinetsu region includes Japan’s capital city, Tokyo, which is the most populous prefecture and also includes Kanagawa, which is the second most populous prefecture. The Kansai region includes Osaka, which is the third most populous prefecture. A large population and the movement of people to and from these regions is thought to have spread the infection, which resulted in medical capacity becoming overwhelmed. Hokkaido, the largest prefecture in Japan, had an increase in the number of patients with COVID-19 in the early phase of the epidemic. Epidemiological evidence to explain this phenomenon remains limited. Before the administration’s state of emergency declaration, the local government in Hokkaido issued its emergency declaration on February 28, 2020, to control the spread of infection [48].

ECMO management in Japanese hospitals has previously resulted in poorer outcomes [49] compared with those reported in other countries during the N1H1 influenza epidemic [39,50-54]. There is a wide consensus that ECMO treatment should be performed at centers with high case volumes and established protocols, and involving clinicians who are experienced in its use. Patients who require ECMO treatment should be transferred to appropriate ECMO centers [39,40,49,54]. Japan has introduced an ECMO project [41] and the ECMOnet [23,25], which involve a consortium working to promote the appropriate use of ECMO, to help develop an evidence-based foundation for its use, to strive for continuous improvement in its application where appropriate, and to collect data concerning the number of patients requiring mechanical ventilation and ECMO support [55,56]. There are >2000 ECMO machines available for use in Japan. The number of patients requiring ECMO was 3.2% of all ECMO machines at the peak of the epidemic, but the capacity levels in terms of use should not be overestimated. In terms of the hospitals participating in the ECMO project in Japan, the number of patients requiring ECMO was 62% of their work at the peak of the epidemic, which is likely to provide a more realistic estimate of their intensive care capacity.

Our study had several limitations. First, the number of critically ill patients requiring intensive care was not completely addressed in the JSICM database. This database has been estimated to capture >80% of clinical cases [25,26]; however, it relies heavily on the cooperation of physicians struggling to treat large numbers of critically ill patients. Second, the extent of the ICU capacity and the number of patients requiring admission to ICUs were not completely validated. The JSICM reported the estimated number of ventilators and ECMO machines according to responses to a questionnaire sent to its member hospitals. Therefore, there was a risk of overestimating ICU capacity. Third, the assumption of one critically ill patient to one depressurized area per hospital did not necessarily reflect the real-life situation. For example, one author’s institution was designated as a Category II infectious diseases institution with eight ICU beds, one depressurized area, and six Category II beds, with one board-certified intensivist. This institution was not a member of the ECMO project. Six Category II beds were fully occupied as soon as the number of infected patients threatened to overwhelm the capacity of the ICU, and other wards were then rearranged to manage patients with mild manifestations of COVID-19. There were two alternatives to treat critically ill patients with COVID-19: one was to use an ICU depressurized bed, and the other was to rearrange the beds in the emergency department. The latter option was chosen because other critically ill and surgical patients could not be treated in the ICU when occupied by patients with COVID-19 for fear of airborne transmissions. Most hospitals are thought to have flexibly employed a variety of methods to increase their capacity to treat patients with COVID-19.

## Conclusions

During the COVID-19 pandemic and following the state of emergency declaration in Japan, the number of patients threatened to overwhelm the number of beds designated for infectious patients; however, Japan’s ICU capacity was sufficient to withstand the demands for mechanical ventilation and ECMO. However, there is no room for complacency in this situation. Urgent consideration is needed concerning future planning concerning how to mitigate the threat to providing intensive care for critically ill patients with COVID-19 at domestic and regional levels in Japan.

## Data Availability

The datasets used and analyzed during the current study are available from the corresponding author on reasonable request.

## Author Contributions

Conceptualization, Y.F. and K.H.; methodology, Y.F.; formal analysis, Y.F.; investigation, Y.F.; data curation, Y.F.; writing—original draft preparation, Y.F.; writing—review and editing, Y.F. and K.H.; visualization, Y.F.; supervision, K.H.; project administration, Y.F. and K.H.; funding acquisition, K.H. All authors have read and agreed to the published version of the manuscript.

## Acknowledgments

The authors would like to thank Editage (www.editage.com) for English language editing.

## Abbreviations

The following abbreviations are used in this manuscript:

WHO: World Health Organization
ECMO: Extracorporeal membranous oxygenation
COVID-19: Coronavirus induced disease 2019
SARS-CoV-2: Severe acute respiratory syndrome coronavirus 2
ICU: Intensive care unit
JSICM: The Japanese Society of Intensive Care Medicine
CRISIS: CRoss Icu Searchable Information System
IQR: Interquartile range

## References

1. World Health Organization. “Coronavirus Disease (COVID-19) Pandemic”. Accessed September 30, 2020. https://www.who.int/emergencies/diseases/novel-coronavirus-2019.

2. Cabinet Secretariat. “Japan’s Response to the Novel Coronavirus Disease: Declaration of a State of Emergency” (April 7, 2020). Accessed September 30, 2020. https://corona.go.jp/en/.

3. Fu L, Wang B, Yuan T, et al. (2020) Clinical characteristics of coronavirus disease 2019 (COVID-19) in China: A systematic review and meta-analysis. J Infect 80:656–665 doi: 10.1016/j.jinf.2020.03.041

4. Chen N, Zhou M, Dong X, et al. (2020) Epidemiological and clinical characteristics of 99 cases of 2019 novel coronavirus pneumonia in Wuhan, China: a descriptive study. Lancet 395:507–513 doi: 10.1016/S0140-6736(20)30211-7

5. Hong X, Xiong J, Feng Z, Shi Y (2020) Extracorporeal membrane oxygenation (ECMO): does it have a role in the treatment of severe COVID-19? Int J Infect Dis 94:78–80 doi: 10.1016/j.ijid.2020.03.058

6. Firstenberg MS, Stahel PF, Hanna J, et al. (2020) Successful COVID-19 rescue therapy by extra-corporeal membrane oxygenation (ECMO) for respiratory failure: a case report. Patient Saf Surg 14:20 doi: 10.1186/s13037-020-00245-7

7. Taniguchi H, Ogawa F, Honzawa H, et al. (2020) Veno-venous extracorporeal membrane oxygenation for severe pneumonia: COVID-19 case in Japan. Acute Med Surg 7:e509 doi: 10.1002/ams2.509

8. MacLaren G, Fisher D, Brodie D (2020) Preparing for the Most Critically Ill Patients With COVID-19: The Potential Role of Extracorporeal Membrane Oxygenation. JAMA doi: 10.1001/jama.2020.2342

9. Hartman ME, Hernandez RA, Patel K, et al. (2020) COVID-19 Respiratory Failure: Targeting Inflammation on VV-ECMO Support. ASAIO J 66:603–606 doi: 10.1097/MAT.0000000000001177

10. Bhimraj A, Morgan RL, Shumaker AH, et al (2020) Infectious Diseases Society of America Guidelines on the Treatment and Management of Patients with COVID-19. Clin Infect Dis. doi: 10.1093/cid/ciaa478

11. Alhazzani W, Møller MH, Arabi YM, et al (2020) Surviving Sepsis Campaign: guidelines on the management of critically ill adults with Coronavirus Disease 2019 (COVID-19). Intensive Care Med 46:854–887. doi: 10.1007/s00134-020-06022-5

12. Beigel JH, Tomashek KM, Dodd LE, et al (2020) Remdesivir for the Treatment of Covid-19 - Preliminary Report. N Engl J Med. doi: 10.1056/NEJMoa2007764

13. Wang Y, Zhang D, Du G, et al (2020) Remdesivir in adults with severe COVID-19: a randomised, double-blind, placebo-controlled, multicentre trial. Lancet 395:1569–1578. doi: 10.1016/S0140-6736(20)31022-9

14. RECOVERY Collaborative Group, Horby P, Lim WS, et al (2020) Dexamethasone in Hospitalized Patients with Covid-19 - Preliminary Report. N Engl J Med. doi: 10.1056/NEJMoa2021436

15. Wang D, Hu B, Hu C, et al. (2020) Clinical Characteristics of 138 Hospitalized Patients With 2019 Novel Coronavirus-Infected Pneumonia in Wuhan, China. JAMA doi: 10.1001/jama.2020.1585

16. Liang W, Liang H, Ou L, et al. (2020) Development and Validation of a Clinical Risk Score to Predict the Occurrence of Critical Illness in Hospitalized Patients With COVID-19. JAMA Intern Med doi: 10.1001/jamainternmed.2020.2033

17. Wang J, Lu F, Zhou M, et al. (2020) Tracheal intubation in patients with severe and critical COVID-19: analysis of 18 cases. Journal of Southern Medical University 40:337–341 doi: 10.12122/j.issn.1673-4254.2020.03.07

18. Sun P, Qie S, Liu Z, et al. (2020) Clinical characteristics of hospitalized patients with SARS-CoV-2 infection: A single arm meta-analysis. J Med Virol 92:612–617 doi: 10.1002/jmv.25735

19. Qiu H, Tong Z, Ma P, et al. (2020) Intensive care during the coronavirus epidemic. Intensive Care Med 46:576–578 doi: 10.1007/s00134-020-05966-y

20. Devereaux A, Christian MD, Dichter JR, et al. (2008) Summary of suggestions from the Task Force for Mass Critical Care summit, January 26-27, 2007. Chest 133:1S–7S doi: 10.1378/chest.08-0649

21. Rubinson L, Hick JL, Curtis JR, et al. (2008) Definitive care for the critically ill during a disaster: medical resources for surge capacity: from a Task Force for Mass Critical Care summit meeting, January 26-27, 2007, Chicago, IL. Chest 133:32S–50S doi: 10.1378/chest.07-2691

22. Li R, Rivers C, Tan Q, et al. (2020) Estimated Demand for US Hospital Inpatient and Intensive Care Unit Beds for Patients With COVID-19 Based on Comparisons With Wuhan and Guangzhou, China. JAMA Netw Open 3:e208297 doi: 10.1001/jamanetworkopen.2020.8297

23. Japan ECMOnet for COVID-19, Shime N (2020) Save the ICU and save lives during the COVID-19 pandemic. J Intensive Care 8:40 doi: 10.1186/s40560-020-00456-1

24. worldometer. “Japan Coronavirus”. Accessed September 30, 2020. https://www.worldometers.info/coronavirus/country/japan/.

25. Japan ECMOnet for COVID-19. “Status of Severe COVID-19 Patients”. Accessed September 30, 2020. https://crisis.ecmonet.jp/.

26. Osamu, N (May 13, 2020). Accessed September 30, 2020. https://www.covid19-jma-medical-expert-meeting.jp/topic/1121. article in Japasene.

27. Japanese Society of Intensive Care Medicine, Society of Respiratory Care Medicine, and Japan Association for Clinical Engineer. “Estimate of the Number of Ventilator, Etc, in Japanese Hospitals”. Accessed September 30, 2020. https://www.jsicm.org/news/news200514.html.

28. Setti L, Passarini F, De Gennaro G, et al. (2020) Airborne Transmission Route of COVID-19: Why 2 Meters/6 Feet of Inter-Personal Distance Could Not Be Enough. Int J Environ Res Public Health 17: doi: 10.3390/ijerph17082932

29. Anderson EL, Turnham P, Griffin JR, Clarke CC (2020) Consideration of the Aerosol Transmission for COVID-19 and Public Health. Risk Analysis 40:902–907 doi: 10.1111/risa.13500

30. Morawska L, Cao J (2020) Airborne transmission of SARS-CoV-2: The world should face the reality. Environ Int 139:105730 doi: 10.1016/j.envint.2020.105730

31. Wilson NM, Norton A, Young FP, Collins DW (2020) Airborne transmission of severe acute respiratory syndrome coronavirus-2 to healthcare workers: a narrative review. Anaesthesia doi: 10.1111/anae.15093

32. Liu Y, Ning Z, Chen Y, et al. (2020) Aerodynamic analysis of SARS-CoV-2 in two Wuhan hospitals. Nature 582:557–560 doi: 10.1038/s41586-020-2271-3

33. Wang J, Feng H, Zhang S, et al. (2020) SARS-CoV-2 RNA detection of hospital isolation wards hygiene monitoring during the Coronavirus Disease 2019 outbreak in a Chinese hospital. Int J Infect Dis 94:103–106 doi: 10.1016/j.ijid.2020.04.024

34. Wax RS, Christian MD (2020) Practical recommendations for critical care and anesthesiology teams caring for novel coronavirus (2019-nCoV) patients. Can J Anaesth 67:568–576 doi: 10.1007/s12630-020-01591-x

35. Ferioli M, Cisternino C, Leo V, et al. (2020) Protecting healthcare workers from SARS-CoV-2 infection: practical indications. Eur Respir Rev 29: doi: 10.1183/16000617.0068-2020

36. Chigurupati R, Panchal N, Henry AM, et al. (2020) Considerations for Oral and Maxillofacial Surgeons in COVID-19 Era: Can We Sustain the Solutions to Keep Our Patients and Healthcare Personnel Safe? Journal of Oral and Maxillofacial Surgery S0278239120305516 doi: 10.1016/j.joms.2020.05.027

37. Huang C, Wang Y, Li X, et al. (2020) Clinical features of patients infected with 2019 novel coronavirus in Wuhan, China. The Lancet 395:497–506 doi: 10.1016/S0140-6736(20)30183-5

38. Ministry of Health, Labour, and Welfare. “Status of Designation of Class II Designated Medical Institutions for Infectious Diseases” (April 1, 2019). Accessed September 30, 2020. https://www.mhlw.go.jp/bunya/kenkou/kekkaku-kansenshou15/02-02-01.html.

39. Haeck JD, Dongelmans DA, Schultz MJ (2012) ECMO Centers and Mortality From Influenza A(H1N1). JAMA 307: doi: 10.1001/jama.2012.57

40. Brodie D, Bacchetta M (2011) Extracorporeal Membrane Oxygenation for ARDS in Adults. N Engl J Med 365:1905–1914 doi: 10.1056/NEJMct1103720

41. Japanese Society of Respiratory Care Medicine. “ECMO Project”. Accessed September 30, 2020. http://square.umin.ac.jp/jrcm/contents/ecmo/index.html.

42. Statistics Bureau of Japan. “Population Statistics on 2019” (October 1, 2019). Accessed September 30, 2020. https://www.stat.go.jp/data/jinsui/2019np/index.html.

43. Zhou F, Yu T, Du R, et al. (2020) Clinical course and risk factors for mortality of adult inpatients with COVID-19 in Wuhan, China: a retrospective cohort study. Lancet 395:1054–1062 doi: 10.1016/S0140-6736(20)30566-3

44. Richardson S, Hirsch JS, Narasimhan M, et al. (2020) Presenting Characteristics, Comorbidities, and Outcomes Among 5700 Patients Hospitalized With COVID-19 in the New York City Area. JAMA doi: 10.1001/jama.2020.6775

45. Guan W-J, Ni Z-Y, Hu Y, et al. (2020) Clinical Characteristics of Coronavirus Disease 2019 in China. N Engl J Med 382:1708–1720 doi: 10.1056/NEJMoa2002032

46. Lois M (2014) The shortage of critical care physicians: is there a solution? J Crit Care 29:1121–1122 doi: 10.1016/j.jcrc.2014.08.001

47. Gutsche JT, Kohl BA (2007) Who should care for intensive care unit patients? Crit Care Med 35:S18–23 doi: 10.1097/01.CCM.0000252907.47050.FE

48. Hokkaido Government. “Declarations a State of Emergency in Hokkaido” (February 28, 2020). Accessed September 30, 2020. http://www.pref.hokkaido.lg.jp/ss/tkk/koronasengen.htm.

49. Takeda S, Kotani T, Nakagawa S, et al. (2012) Extracorporeal membrane oxygenation for 2009 influenza A(H1N1) severe respiratory failure in Japan. J Anesth 26:650–657 doi: 10.1007/s00540-012-1402-x

50. Checkley W (2011) Extracorporeal Membrane Oxygenation as a First-Line Treatment Strategy for ARDS: Is the Evidence Sufficiently Strong? JAMA 306:1703 doi: 10.1001/jama.2011.1504

51. Noah MA, Peek GJ, Finney SJ, et al. (2011) Referral to an extracorporeal membrane oxygenation center and mortality among patients with severe 2009 influenza A(H1N1). JAMA 306:1659–1668 doi: 10.1001/jama.2011.1471

52. Laudi S, Busch T, Kaisers U (2010) Extracorporeal membrane oxygenation for ARDS due to 2009 influenza A(H1N1). JAMA 303:941; author reply 942 doi: 10.1001/jama.2010.200

53. Australia and New Zealand Extracorporeal Membrane Oxygenation (ANZ ECMO) Influenza Investigators, Davies A, Jones D, et al. (2009) Extracorporeal Membrane Oxygenation for 2009 Influenza A(H1N1) Acute Respiratory Distress Syndrome. JAMA 302:1888–1895 doi: 10.1001/jama.2009.1535

54. Peek GJ, Mugford M, Tiruvoipati R, et al. (2009) Efficacy and economic assessment of conventional ventilatory support versus extracorporeal membrane oxygenation for severe adult respiratory failure (CESAR): a multicentre randomised controlled trial. Lancet 374:1351–1363 doi: 10.1016/S0140-6736(09)61069-2

55. Japan ECMOnet for COVID-19 (2020) Japan ECMOnet for COVID-19: telephone consultations for cases with severe respiratory failure caused by COVID-19. J Intensive Care 8:24 doi: 10.1186/s40560-020-00440-9

56. Japan ECMOnet for COVID-19 (2020) Nationwide system to centralize decisions around ECMO use for severe COVID-19 pneumonia in Japan (Special Correspondence). J Intensive Care 8:29 doi: 10.1186/s40560-020-00445-4

